# Factors associated with correct vaccination among children aged 24 to 48 months hospitalized in the national hospitals of Bujumbura

**DOI:** 10.1101/2024.03.18.24304508

**Authors:** Felix Niyongabo, Jean M. Butoyi, Niyoyunguruza Laetitia, Kabusoni Esperance, Yves Coppieters

**Affiliations:** East African Nutritional Sciences Institute, University of Burundi, Burundi; Faculty of Health Sciences, Lumière University of Bujumbura, Burundi; Department of Paraclinical Sciences National Institute of Public Health, Bujumbura, Burundi; Health Systems and Policies – International Health, School of Public Health, Université Libre de Bruxelles (ULB), Brussels, Belgium

**Keywords:** associated factors, correct vaccination, national hospitals, children aged 24 to 48 months

## Abstract

**Introduction:** In Africa, although vaccines are often available free of charge in health facilities, many children evade various strategies put in place to reach them. In recent years, despite Burundi maintaining high coverage in all antigens (over 80% since the launch of the PEV in 1980), performance has begun to decline. From 2015 to 2017, all coverage rates experienced a slight regression, and the WHO/UNICEF figures for 2018 confirm this downward trend. The objective of this work is to contribute to the reduction of infant morbidity and mortality due to vaccine-preventable diseases by identifying factors associated with correct vaccination among children aged 24 to 48 months hospitalized in national hospitals in Bujumbura.

**Methodology:** A cross-sectional analytical study was conducted in 4 national hospitals in Burundi. A sample of 216 children was selected, and the collected data were entered into Epinfo 7.2 software and transferred to Stata 15 for analysis. Three types of analyses were performed: descriptive analysis, univariate logistic regression, and multivariate logistic regression. After modeling, variables with a p-value <0.05 were considered associated with correct vaccination.

**Results:** Out of 216 children, correct vaccination was observed in 64.81%. The average age of mothers was 29.4 ± 5.43 years, ranging from 15 to 45 years. Factors significantly associated with correct vaccination after modeling were: husband’s level of education [OR = 15.41; P = 0.021], household income [OR = 10.23; P = 0.021], distance from residence to vaccination facility [OR = 0.12; P = 0.000], and mothers’ knowledge levels of vaccine-preventable diseases [OR = 1.73; P = 0.004]. The model was well-calibrated (the p-value of the Lemeshow test is 0.6697) with a 90% capacity to correctly classify observations.

**Conclusion:** The study’s results indicate that children are not properly vaccinated despite the service being free. Various factors influencing correct vaccination status were identified, including the level of education of the head of the household, income level, distance to vaccination facilities, and mothers’ knowledge of vaccine-preventable diseases. It is essential to implement interventions targeting these factors associated with correct vaccination to reduce morbidity and mortality from vaccine-preventable diseases.

## I. Introduction

### I.1. General Context

Vaccination is one of the effective public health measures to prevent mortality and infectious complications among children worldwide. It is also an indicator of development in most low- and middle-income countries. According to the WHO, nearly 3 million deaths are prevented annually worldwide through vaccination, and 1.5 million additional deaths could be avoided simply by improving vaccination status, at a time when 19.5 million infants worldwide are not adequately vaccinated. By controlling deadly or disabling communicable diseases, vaccination remains one of the essential interventions for achieving sustainable development goals. In most developed countries, there are still groups of individuals who are incompletely vaccinated. Recent studies show that in the United States, one out of eight children under 2 years old is not properly vaccinated, while in Italy, there has been a decline in vaccination coverage rates for various vaccine-preventable diseases since 2013, dropping from 95.7% in 2013 to 93.3% in 2016. In Africa, although vaccination services are often available free of charge in health facilities, many children evade various strategies implemented to reach them, leading to high rates of missed vaccination opportunities.

### I.2. Specific Context

In Burundi, according to the DHS 2016-2017, only 85% of children aged 12-23 months have received all basic vaccines, 72% have received all vaccines for the appropriate age group, while a small proportion of children (0.3%) have received no vaccines. Vaccination coverage varies according to certain sociodemographic characteristics; the proportion of children receiving all basic vaccines is higher in rural areas than in urban areas (86% versus 80%). There are also losses between vaccine doses concerning Pentavalent, Polio, and pneumococcal vaccines, with a 99% loss for the first dose of Pentavalent, decreasing to 97% for the third dose. For Polio, the loss is more significant, dropping from 99% for the first dose to 92% for the third dose. Regarding pneumococcal vaccines, between the first and third doses, the proportion decreases from 98% to 94%. Despite Burundi maintaining satisfactory coverage for all antigens (over 80% since the launch of the PEV in 1980), performance in recent years has begun to decline. From 2015 to 2017, all coverage rates experienced a slight regression, and the WHO/UNICEF figures for 2018 confirm this downward trend. The results of the coverage and equity survey conducted in 2018 showed that these underperformances varied from one district to another.

### I.3. Research Questions

What are the explanatory factors for correct vaccination status among children hospitalized in the four national hospitals of Burundi?

### I.4. Objectives

#### I.4.1. **General Objective**

To contribute to the reduction of infant morbidity and mortality due to vaccine-preventable diseases by identifying factors associated with correct vaccination among children aged 24 to 48 months hospitalized in national hospitals in Bujumbura.

#### I.4.2. Specific Objectives

- To explore the association between sociodemographic characteristics (mother’s age, level of education, marital status, religion) and correct vaccination status among children aged 24 to 48 months hospitalized in the national hospitals of Bujumbura municipality.
- To study the association between child characteristics (gender, birth order) aged 24 to 48 months and correct vaccination status in the national hospitals of Bujumbura municipality.
- To investigate the existence of an association between correct vaccination status and accessibility to healthcare facilities among children aged 24 to 48 months hospitalized in the national hospitals of Bujumbura municipality.
- To identify household characteristics (standard of living, household size) associated with correct vaccination status for children aged 24 to 48 months hospitalized in the national hospitals of Bujumbura municipality.
- Achieving the study’s objectives will enable the formulation of recommendations to the health district teams of the Bujumbura municipality and the Ministry of Public Health and the Fight against AIDS regarding the implementation of activities aimed at promoting infant vaccination.

### I.5. Research Hypotheses

- Child characteristics (gender, birth order) are associated with infant vaccination completeness.
- Household characteristics (standard of living, household size) are associated with correct infant vaccination.
- Maternal characteristics (mother’s age, level of education, etc.) are associated with correct infant vaccination.
- Accessibility to vaccination facilities (distance from residence to vaccination facility) is associated with correct vaccination among infants.

## II. Methods

### II.1. Study Population

Our study population consisted of infants aged 24 to 48 months hospitalized in the four national hospitals of Bujumbura municipality (Prince Régent Charles Hospital, Prince Louis Rwagasore Clinic, Kamenge Military Hospital, Kamenge University Hospital Center).

### II.2. Study Type

A cross-sectional analytical study was conducted. The survey was conducted among mothers or caregivers of children aged 24 to 48 months hospitalized in the four national hospitals of Burundi.

### II.3. Study Period

The study was conducted over a period of 1 month from December 2019 to January 2020 in the 4 national hospitals of Burundi.

### II.4. Sampling

#### II.4.1. **Sampling Technique**

A comprehensive sampling of all children aged 24 to 48 months hospitalized in the four national hospitals of Bujumbura municipality during the study period was conducted.

### II.4.2. Sample Size

The minimum sample size was estimated using the following formula:

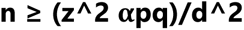

Where:

n: sample size

p: proportion of children aged 13 to 23 months with incorrect vaccination status. To estimate this proportion, we considered the proportion of children who did not complete their vaccination schedule before their first birthday, which is 14.9%, according to the DHS 2016-2017.

q: proportion of children aged 12 to 23 months with optimal vaccination status

q = 1 - p = 1 - 0.149 = 0.851

d: desired absolute precision level (d = 0.05)

z*α*: Confidence coefficient (at a significance level of 0.05)

In anticipation of a non-response rate of up to 10%, the minimum sample size was estimated at 216.

#### II.4.3. Data Collection Tools

Data were collected in two ways based on a questionnaire:

From vaccination cards for information related to vaccination status (which allowed for the determination of vaccination coverage and vaccination schedules),

From responses provided by the mother/caregiver for sociodemographic characteristics of mothers/caregivers and households, child characteristics, and accessibility to vaccination centers. Informed consent has been obtained beforehand from parents or guardians and the author had access to information that could identify individual participants during data collection.

#### II.4.4. Inclusion Criteria

Infants hospitalized in one of the four national hospitals of Bujumbura Municipality aged between 24 and 48 months during the survey period, whose mothers or caregivers consented to participate in the study, were included in this study.

#### II.4.5. Exclusion Criteria

Infants aged 24 to 48 months whose mothers were unavailable and whose caregivers were unable to provide any vaccination-related responses or for whom the vaccination card was not available were excluded from the study.

### II.6. Data Analysis

Data entry was performed using Epi Info software, then transferred to Stata 15 for analysis. Three types of analyses were conducted:

#### II.6.1. Descriptive Analysis

Weighted proportions or percentages were calculated for qualitative variables, mean and standard deviation for normally distributed quantitative variables. Median and interquartile range (Q1 and Q3) were calculated for non-normally distributed quantitative variables.

#### II.6.2. Bivariate Analysis

For the search for an association between children’s vaccination status and various factors, bivariate analysis of the data was performed using the Pearson Chi-square test. The associations between the dependent variable and the independent variables were measured by odds ratios (OR) and their 95% confidence intervals (CI).

#### II.6.3. Multivariate Analysis

A logistic regression model was used to identify independent variables associated with correct vaccination status of children. All independent variables with significance levels less than or equal to 20% in bivariate analysis were included in the initial model. Stepwise modeling was then performed to determine statistically significant variables using the backward variable selection method. The Akaike Information Criterion (AIC) was used to select the best model (one with a low AIC), and a final model was retained. The significance level was set at 5%.

### II.7. Study Validity

#### II.7.1. Internal Validity

To mitigate biases, medical personnel were recruited as investigators and trained on the survey objectives and data collection procedures. The questionnaire was tested one week before use at Prince Régent Charles Hospital. Selection bias was circumvented by exhaustive sampling.

#### II.7.2. External Validity

The study results will be inferred to the population of children aged 24 to 48 months hospitalized in the national hospitals of Burundi.

### II.8. Ethical Aspects and Protection of Personal Data

The research protocol received approval from the INSP ethics committee. Permissions were obtained from the medical directors of the four national hospitals before conducting the survey. Prior to administering the questionnaire, oral informed consent was obtained. Data collection was anonymous. Access to filled survey forms was limited to the research team members only. The forms were kept in a locked cabinet before and after data entry. Team members are bound by confidentiality for all information collected during the study.

Participation in this study carries no risk for respondents. They were free to refuse to participate in the study or to answer any questions deemed intrusive without any consequences.

The study also does not offer any direct benefits to respondents, but their participation will contribute to the adaptation of vaccination implementation approaches and maximize their health benefits for children.

## III. Results

The presentation of the results revolves mainly around three points:

- Description of the sample
- Bivariate analysis
- Multivariate analysis

### A. Description of the Sample

Information regarding the description of the sample is provided according to the following characteristics: sociodemographic characteristics of the mother and accessibility to healthcare facilities, characteristics of the children, as well as mothers’ knowledge of the vaccination schedule and vaccine-preventable diseases.

#### 1. Sociodemographic Characteristics of the Mother

The mean age of the mothers is 29.4 ± 5.43 years, with an age range from 15 to 45 years.

**Table 3:** Sociodemographic Characteristics of the Mother and Access to Healthcare Facilities.

| **Characteristics** | **Frequency** | **Percentage (%)** |
| --- | --- | --- |
| **Age** |  |  |
| 15 years - 25 years | 54 | 25.4% |
| 25 years - 35 years | 134 | 62.0% |
| 35 years - 45 years | 27 | 12.5% |
| **Mother's Education Level** |  |  |
| None | 36 | 14.3% |
| Primary | 66 | 30.5% |
| Secondary | 73 | 33.8% |
| Higher Education | 46 | 21.3% |
| **Marital Status** |  |  |
| Married | 193 | 89.3% |
| Single | 11 | 5.0% |
| Widowed | 11 | 5.0% |
| Divorced/Separated | 1 | 0.4% |
| **Father's Education Level** |  |  |
| None | 13 | 6.0% |
| Primary | 52 | 24.0% |
| Secondary | 66 | 30.5% |
| Higher Education | 62 | 28.7% |
| Unknown | 23 | 10.6% |
| **Religion** |  |  |
| Catholic | 83 | 38.4% |
| Protestant | 90 | 41.6% |
| Jehovah's Witnesses | 2 | 0.9% |
| Muslim | 41 | 18.9% |
| **Standard of Living** |  |  |
| Low | 85 | 39.3% |
| Medium | 108 | 50.0% |
| High | 23 | 10.6% |
| **Household Size** |  |  |
| 2-5 Individuals | 158 | 73.1% |
| More than 5 Individuals | 58 | 26.8% |
| **Distance** |  |  |
| Less than 5KM | 137 | 63.4% |
| More than 5KM | 79 | 36.5% |

The age group of 25-35 years was the most represented, accounting for 62.04%. The average age was 29 years, ranging from 15 to 45 years. Regarding the educational level, 30.5% of mothers had a primary education, while 33.8% had a secondary education. Married women were the majority at 89.3%. Secondary and higher education levels among husbands were represented by 30.5% and 28.7%, respectively. Children whose mothers belonged to the Protestant religion dominated at 41.6%, while the Catholic religion was represented at 38.4%. Children from families with a medium standard of living represented half of the population. A household size of 2 to 5 individuals was found in 73.1% of observations, while in 63.4% of cases, the distance from home to the vaccination facility was less than 5 kilometers

#### 1. Child Characteristics

The table below highlights various characteristics of the children, including vaccination status, gender, and the child’s birth order in the sibling hierarchy.

**Table 4:**
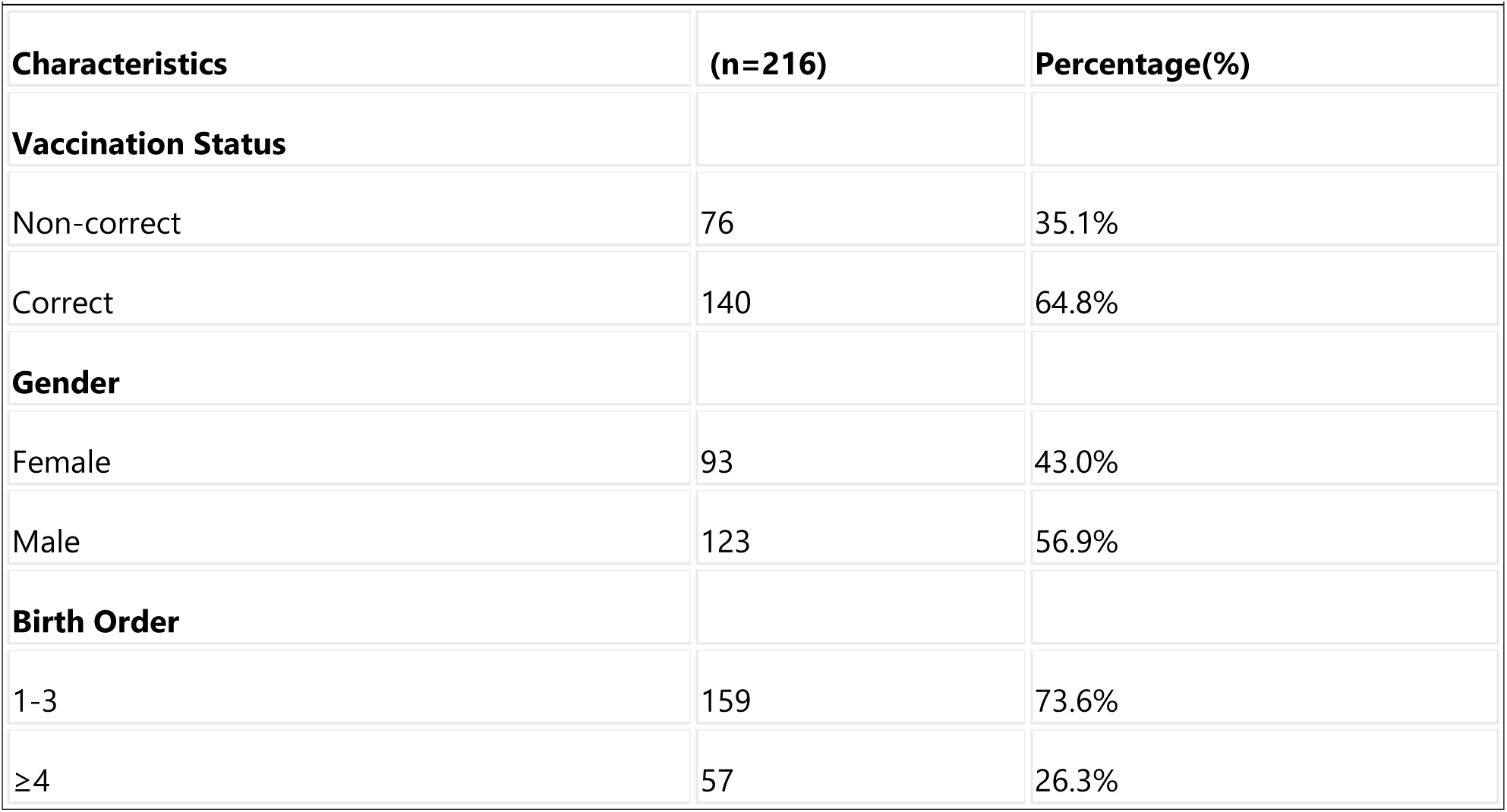
Child Characteristics.

The correct vaccination rate was observed in 64.81%. Male children accounted for 56.94%, while the birth order ranging from the first to the third child was associated with 73.61% of the observations.

#### 2. Mothers’ Knowledge of Vaccination Schedule and Vaccine-Preventable Diseases (VPDs)

Below is the table illustrating the level of mothers’ knowledge about the vaccination schedule and vaccine-preventable diseases.

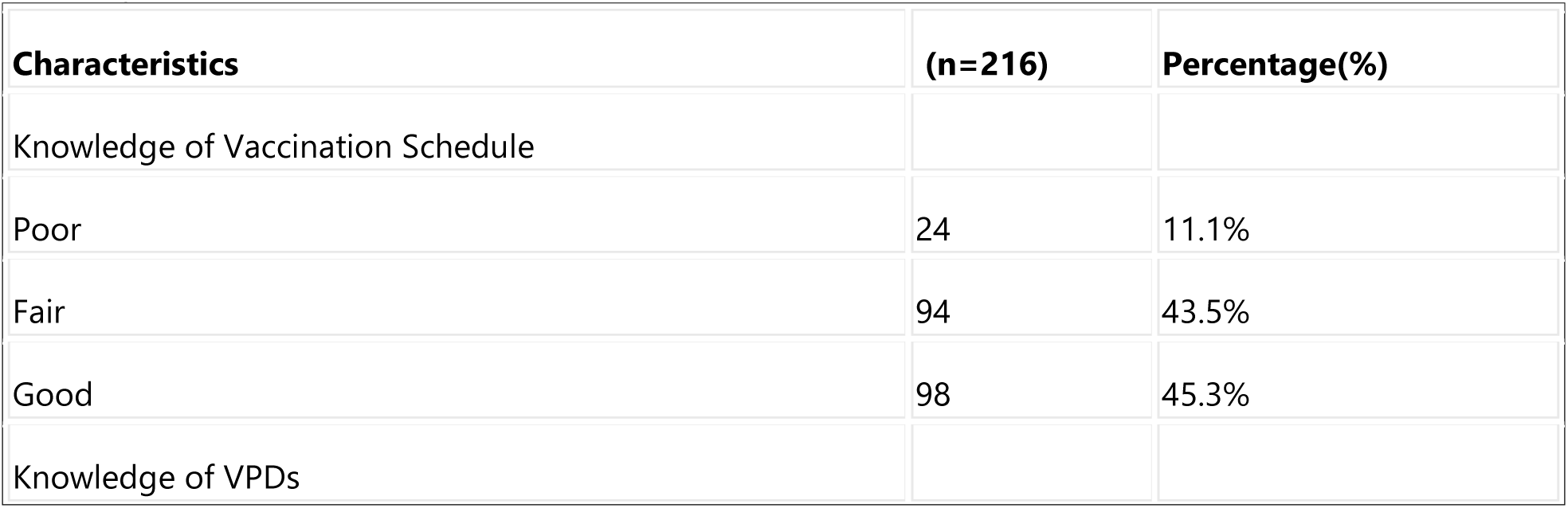
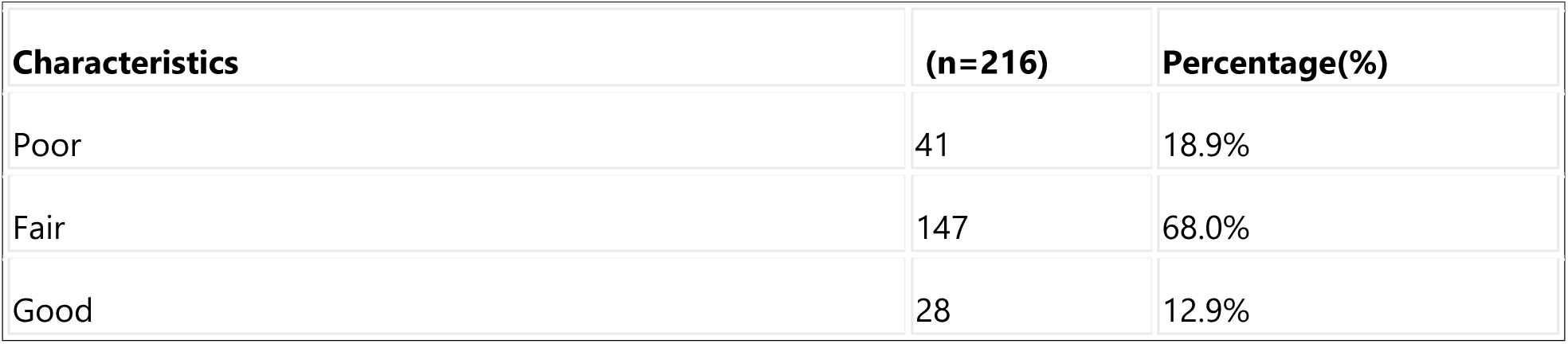

| **Characteristics** | **(n=216)** | **Percentage(%)** |
| --- | --- | --- |
| **Knowledge of Vaccination Schedule** |  |  |
| Poor | 24 | 11.1% |
| Fair | 94 | 43.5% |
| Good | 98 | 45.3% |
| **Knowledge of VPDs** |  |  |

| Characteristics | (n=216) | Percentage(%) |
| --- | --- | --- |
| Poor | 41 | 18.9% |
| Fair | 147 | 68.0% |
| Good | 28 | 12.9% |

This table illustrates that 11.11% of mothers had poor knowledge of the vaccination schedule, while 18.98% had poor knowledge of vaccine-preventable diseases.

### A. Bivariate Analysis

To test the independence between the correct vaccination status and the various variables in our study, a cross-tabulation of the correct vaccination status variable with other variables was performed using the Chi-square test.

#### B.1. Investigation of Relationship between Correct Vaccination Status of Children and Various Factors using Pearson’s Chi-Square Test

##### B.1.1. Correct Vaccination Status based on Sociodemographic Characteristics of the Mother and Access to Healthcare Facilities

**Table 6:**
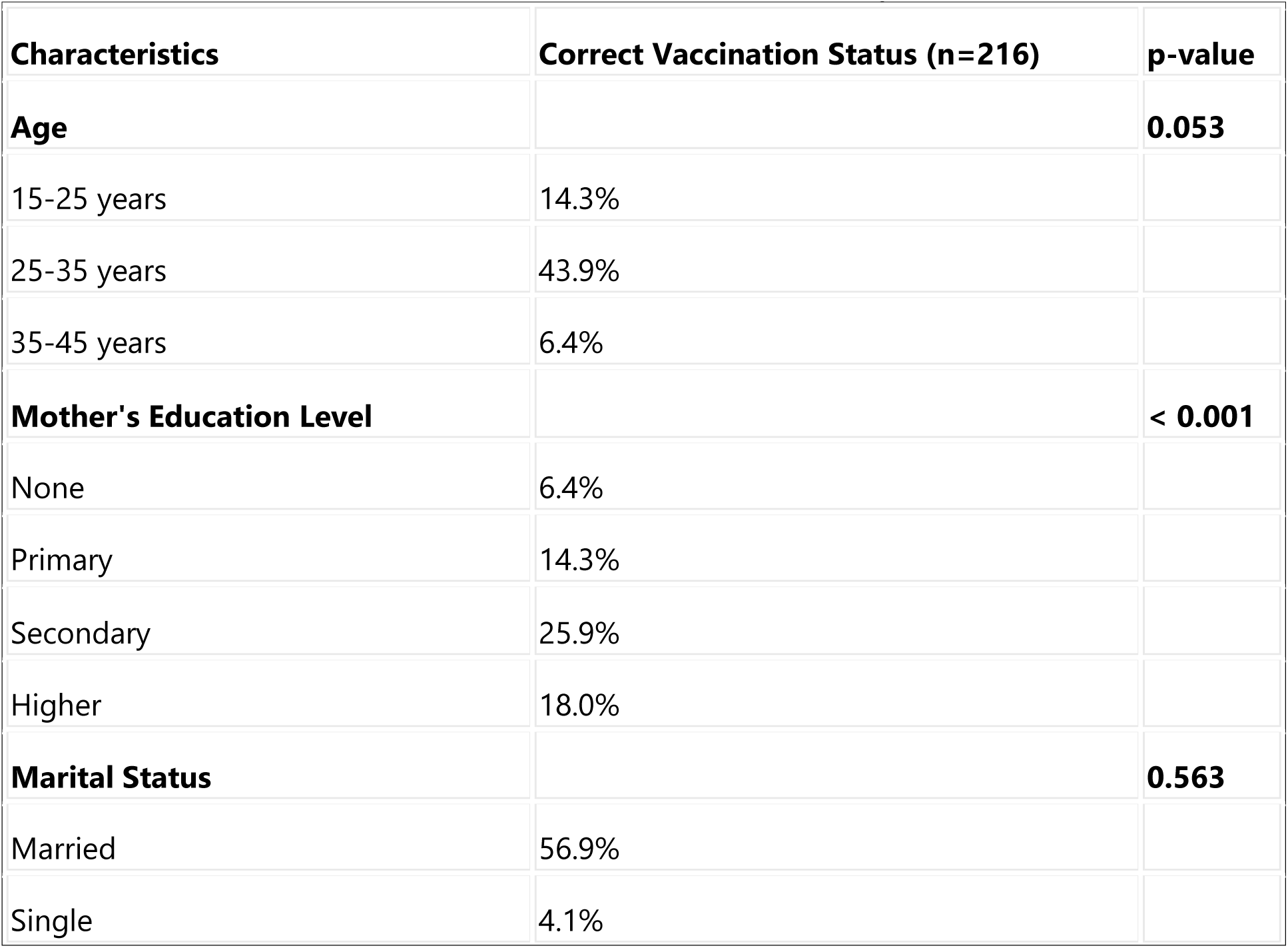
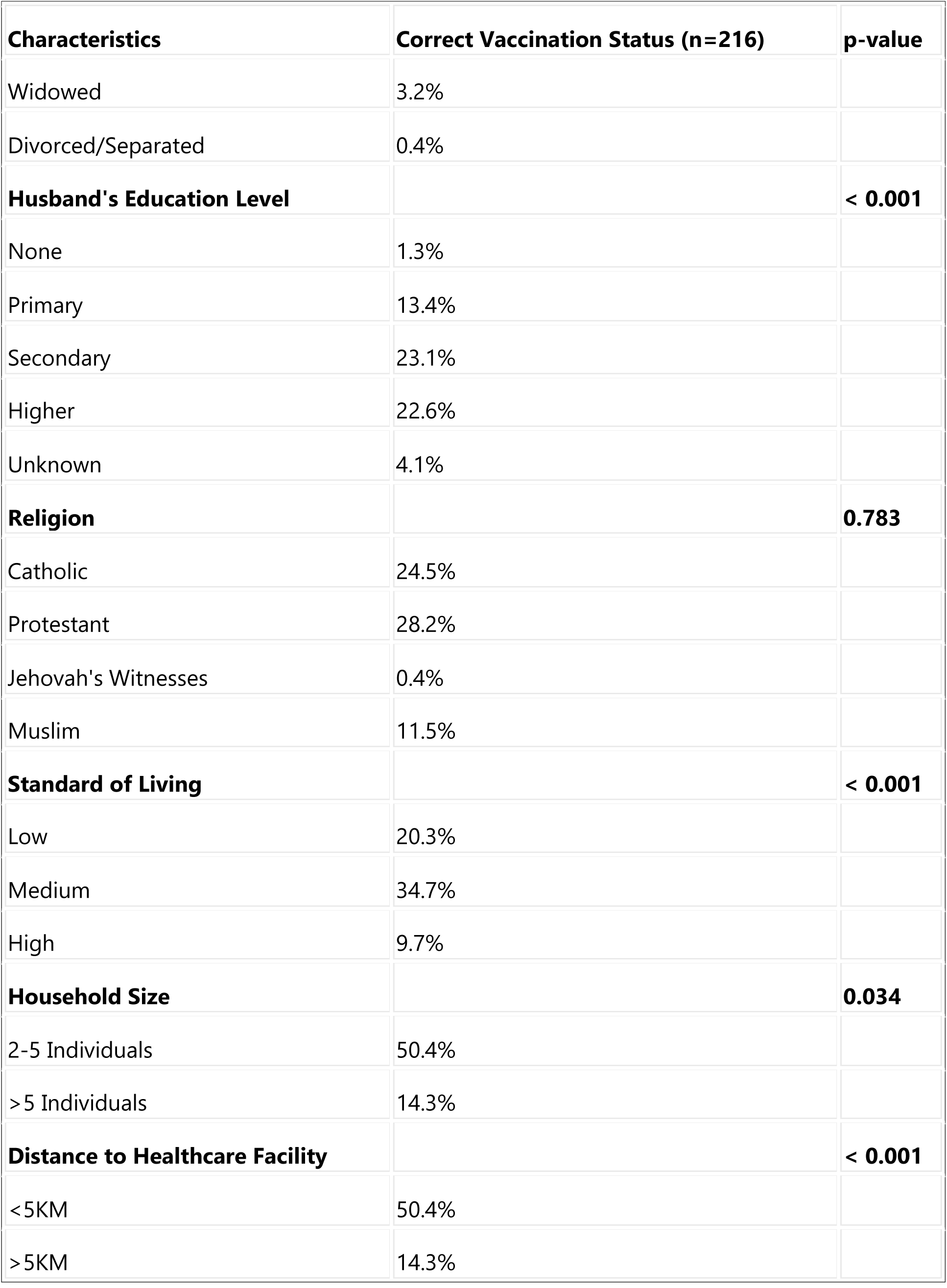
Relationship between Correct Vaccination Status and Sociodemographic Characteristics of the Mother and Access to Healthcare Facility.

##### B.1.2. Correct Vaccination Status based on Child Characteristics

The table below highlights the results of the independence test between correct vaccination status and child characteristics.

**Table 7:** Relationship between Correct Vaccination Status and Child Characteristics.

| Characteristics | Correct Vaccination Status | p-value |
| --- | --- | --- |
| **Sex** |  | **0.129** |
| Female | 25.4% |  |
| Male | 39.3% |  |
| **Birth Order** |  | **0.098** |
| 1-3 | 50.0% |  |
| 4-6 | 14.3% |  |
| ≥7 | 0.4% |  |

##### B.1.3. Correct Vaccination Status based on Mothers’ Knowledge of Vaccination Schedule and Vaccine-Preventable Diseases (VPDs)

The table below shows the results of the independence test between correct vaccination status and the level of mothers’ knowledge about the vaccination schedule and vaccine-preventable diseases.

**Table 8:** Relationship between Correct Vaccination Status and Mothers’ Knowledge of Vaccination Schedule and VPDs.

| Characteristics | Correct Vaccination Status (n=216) | p-value |
| --- | --- | --- |
| **Knowledge of Vaccination Schedule** |  | **< 0.001** |
| Poor | 21.7% |  |
| Fair | 8.3% |  |
| Good | 34.7% |  |
| **Knowledge of VPDs** |  | **< 0.001** |
| Poor | 36.1% |  |
| Fair | 16.2% |  |
| Good | 12.5% |  |

For this part, the relationships between explanatory factors and the vaccination variable were tested using the Chi-square statistical test, and the results were recorded in the above table.

The variables for which a statistically significant difference was found compared to the study variable are: mother’s education level, mother’s husband’s education level, household standard of living, household size, distance between home and vaccination location, knowledge of vaccination schedule, and knowledge of vaccine-preventable diseases. However, no significant relationship was found between the child’s vaccination status and the mother’s age, marital status, mother’s religion, child’s gender, and birth order in the sibling hierarchy.

#### B.2. Measurement of Association between Correct Vaccination Status and Independent Variables

Univariate logistic regression was used to determine potential predictors of correct vaccination status. Odds ratios were estimated for all variables with a statistically significant association with correct vaccination status, and the results are presented in Tables 10, 11, and 12 below

**Table 9:**
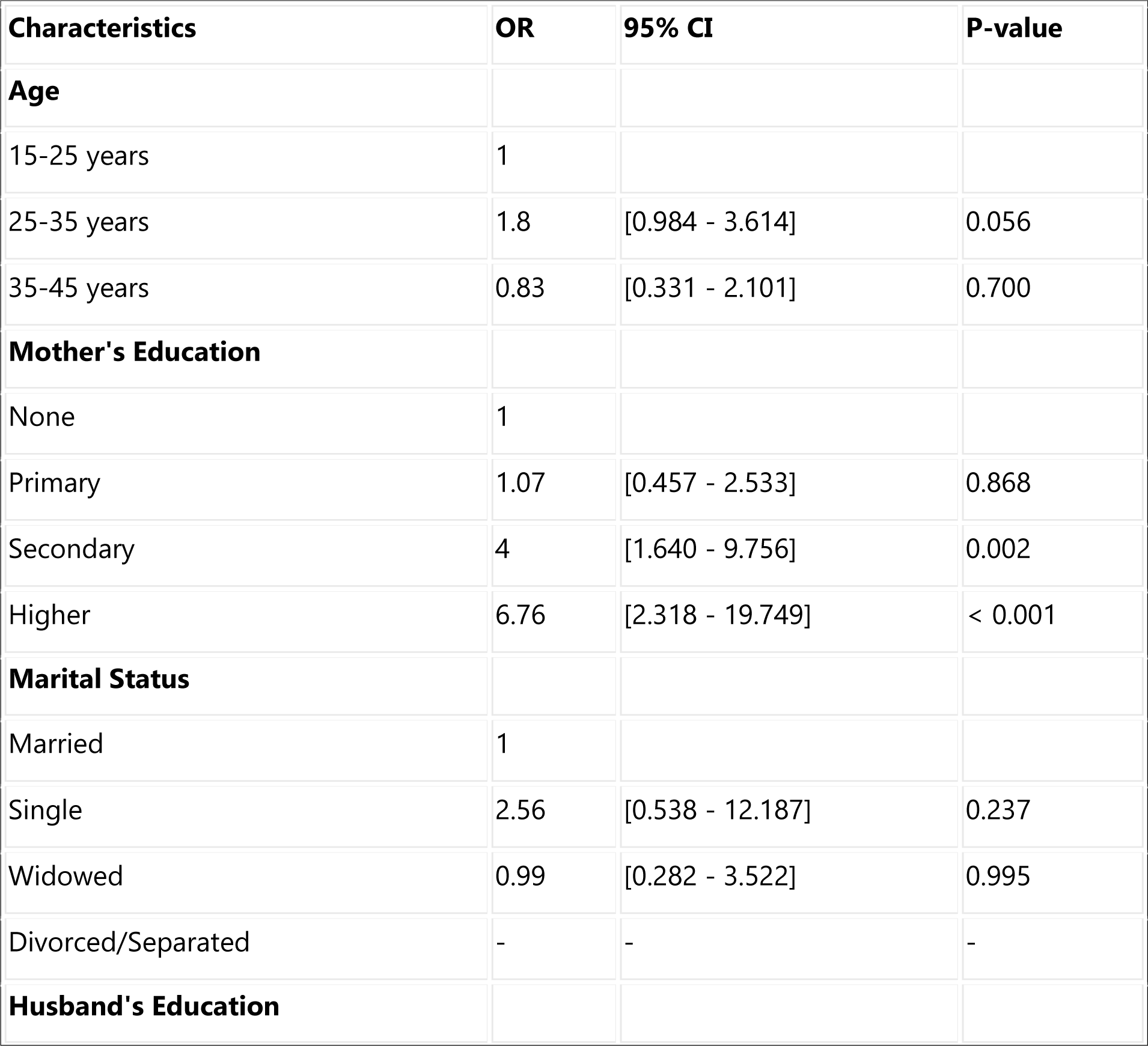
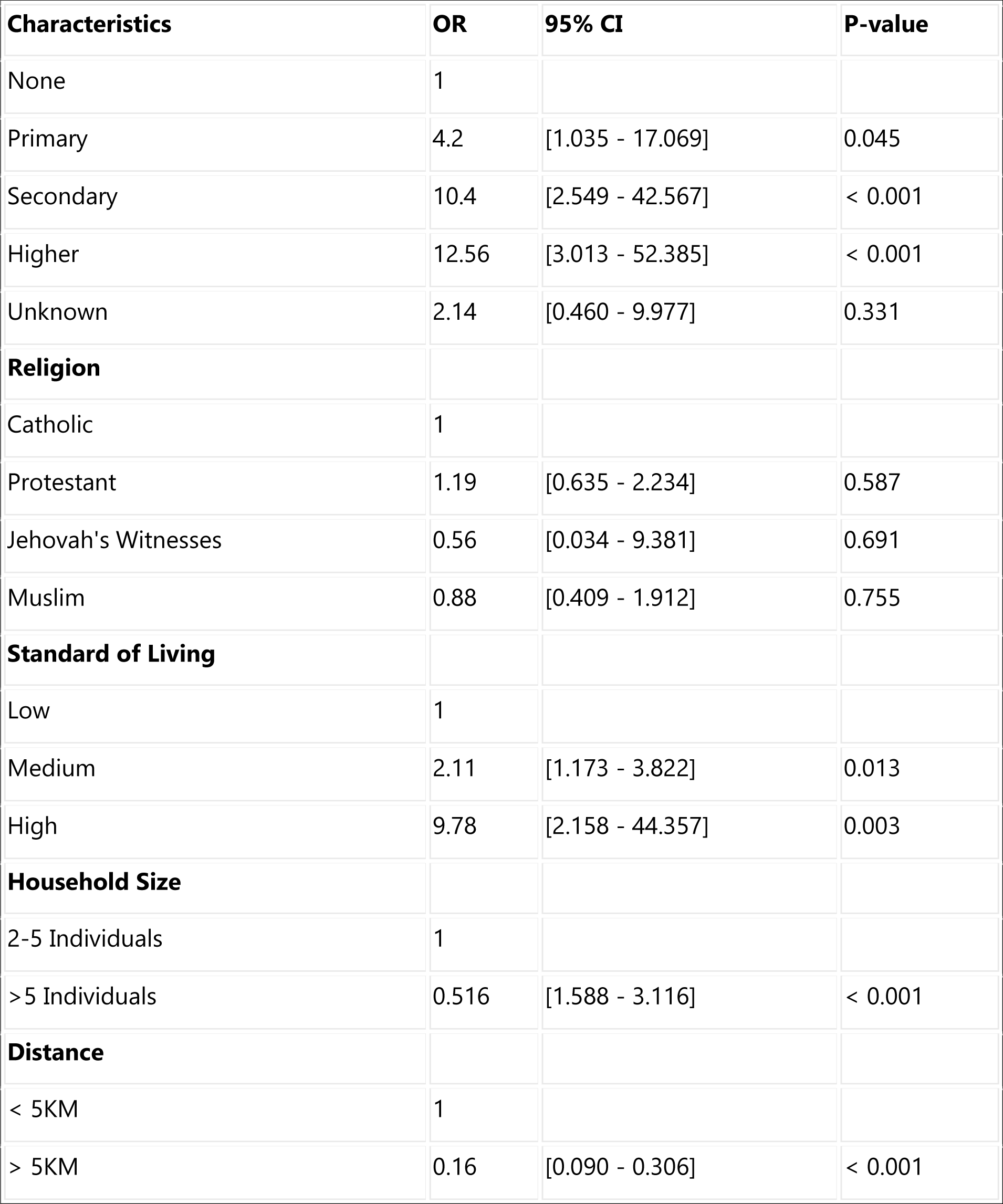
Association between Correct Vaccination Status and Sociodemographic Characteristics of the Mother and Access to Healthcare Facility.

| **Characteristics** | **OR** | **95% CI** | **P-value** |
| --- | --- | --- | --- |
| **Age** |  |  |  |
| 15-25 years | 1 |  |  |
| 25-35 years | 1.8 | [0.984 - 3.614] | 0.056 |
| 35-45 years | 0.83 | [0.331 - 2.101] | 0.700 |
| **Mother's Education** |  |  |  |
| None | 1 |  |  |
| Primary | 1.07 | [0.457 - 2.533] | 0.868 |
| Secondary | 4 | [1.640 - 9.756] | 0.002 |
| Higher | 6.76 | [2.318 - 19.749] | < 0.001 |
| **Marital Status** |  |  |  |
| Married | 1 |  |  |
| Single | 2.56 | [0.538 - 12.187] | 0.237 |
| Widowed | 0.99 | [0.282 - 3.522] | 0.995 |
| Divorced/Separated | - | - | - |
| **Husband's Education** |  |  |  |
| None | 1 |  |  |
| Primary | 4.2 | [1.035 - 17.069] | 0.045 |
| Secondary | 10.4 | [2.549 - 42.567] | < 0.001 |
| Higher | 12.56 | [3.013 - 52.385] | < 0.001 |
| Unknown | 2.14 | [0.460 - 9.977] | 0.331 |
| **Religion** |  |  |  |
| Catholic | 1 |  |  |
| Protestant | 1.19 | [0.635 - 2.234] | 0.587 |
| Jehovah's Witnesses | 0.56 | [0.034 - 9.381] | 0.691 |
| Muslim | 0.88 | [0.409 - 1.912] | 0.755 |
| **Standard of Living** |  |  |  |
| Low | 1 |  |  |
| Medium | 2.11 | [1.173 - 3.822] | 0.013 |
| High | 9.78 | [2.158 - 44.357] | 0.003 |
| **Household Size** |  |  |  |
| 2-5 Individuals | 1 |  |  |
| >5 Individuals | 0.516 | [1.588 - 3.116] | < 0.001 |
| **Distance** |  |  |  |
| < 5KM | 1 |  |  |
| > 5KM | 0.16 | [0.090 - 0.306] | < 0.001 |

**Table 10:** Association between Vaccination Status and Child Characteristics.

| Characteristics | OR | 95% CI | P-value |
| --- | --- | --- | --- |
| **Sex** |  |  |  |
| Female | 1 |  |  |
| Male | 1.54 | [.880 - 2.714] | 0.130 |
| **Birth Order** |  |  |  |
| 1-3 | 1 |  |  |
| ≥ 4 | 0.60 | [.325 - 1.124] | 0.112 |

**Table 11:** Association between Vaccination Status and Mothers’ Knowledge of Vaccination Schedule and VPDs.

| Characteristics | OR | 95% CI | P-value |
| --- | --- | --- | --- |
| **Knowledge of Vaccination Schedule** |  |  |  |
| Poor | 1 |  |  |
| Fair | 0.33 | [0.122 - 0.914] | 0.033 |
| Good | 1.08 | [0.386 - 3.061] | 0.875 |
| **Knowledge of VPDs** |  |  |  |
| Poor | 1 |  |  |
| Fair | 1.91 | [0.077 - 0.489] | < 0.001 |
| Good | 4.62 | [0.525 - 40.771] | 0.167 |

**Table 12:** Multivariate Analysis (Final Logistic Model)

| Explanatory Variables | Adjusted OR | 95% CI | P-value |
| --- | --- | --- | --- |
| **AGE (Reference: 15-25 years)** | **1** |  |  |
| 25-35 years | 0.65 | [0.22 - 1.86] | 0.424 |
| 35-45 years | 0.34 | [0.07 - 1.62] | 0.178 |
| **Mother's Education (Reference: None)** | **1** |  |  |
| Primary | 0.38 | [0.10 - 1.39] | 0.145 |
| Secondary | 2.76 | [0.54 - 14.03] | 0.221 |
| Higher | 1.38 | [0.14 - 13.70] | 0.780 |
| **Marital Status (Reference: Married)** | **1** |  |  |

| **Explanatory Variables** | **Adjusted OR** | **95% CI** | **P-value** |
| --- | --- | --- | --- |
| Single | 5.05 | [0.47 - 53.75] | 0.179 |
| Widowed | 1.98 | [0.24 - 16.30] | 0.522 |
| **Husband's Education (Reference: None)** | **1** |  |  |
| Primary | 15.41 | [1.52 - 156.15] | 0.021 |
| Secondary | 10.98 | [0.91 - 131.22] | 0.058 |
| Higher | 9.30 | [0.55 - 155.27] | 0.120 |
| Unknown | 2.40 | [0.17 - 33.36] | 0.512 |
| **Standard of Living (Reference: Low)** | **1** |  |  |
| Medium | 2.43 | [0.94 - 6.28] | 0.066 |
| High | 10.23 | [1.41 - 74.14] | 0.021 |
| **Household Size (Reference: 2-5)** | **1** |  |  |
| >5 | 0.50 | [0.17 - 1.40] | 0.191 |
| **Distance (Reference: ≤5km)** | **1** |  |  |
| >5km | 0.12 | [0.03 - 0.24] | < 0.001 |
| Sex (Reference: Female) | 1 |  |  |
| Male | 1.3 | [0.56 - 3.13] | 0.507 |
| **Knowledge of Vaccination Schedule (Reference: Poor)** | **1** |  |  |
| Fair | 0.65 | [0.14 - 2.89] | 0.574 |
| Good | 1.62 | [0.38 - 6.94] | 0.509 |
| **Knowledge of VPDs (Reference: Poor)** |  |  |  |
| Fair | 1.73 | [0.03 - 0.51] | 0.004 |
| Good | 4.92 | [0.27 - 89.36] | 0.281 |

From the above table, the association of maternal and child characteristics and access to a healthcare facility with the vaccination status of children was evaluated using bivariate analysis. In this study, the mother’s education was statistically associated with correct vaccination at the bivariate level. Mothers with a secondary education were 4 times more likely [Odd Ratio (OR) = 4] to vaccinate their children correctly than mothers with no education, while those with a higher education were 7 times more likely [OR = 6.76] to vaccinate their children correctly than mothers with no education.

The father’s education level also showed a positive influence on the likelihood of the child being correctly vaccinated. Children born to fathers with a primary education were 4 times more likely [OR = 4.2] to be correctly vaccinated than children born to fathers with no education. Those born to fathers with secondary and higher levels of education were respectively 10 times [OR = 10.4] and 12 times [OR = 12.56] more likely to be correctly vaccinated than children born to fathers with no education.

It was also observed that the household’s standard of living had a positive association with complete vaccination of children. Children from households with a medium standard of living were 2 times more likely to have correct vaccination [OR = 2.11] compared to those from households with a low standard of living, while those from households with a high standard of living were 10 times more likely to have complete vaccination [OR = 9.78] compared to those from households with a low standard of living. For household size, children from households with more than 5 individuals were 0.5 times more likely [OR = 0.516] to complete vaccination compared to children from households with 2 to 5 individuals.

The distance to the healthcare facility influences a child’s vaccination status. Children born to mothers who report that the distance from their home to the vaccination center is a problem were 0.16 times likely to receive complete vaccination than children of mothers who report that distance is not a problem [OR = 0.16].

##### B.2.2. Vaccination Status and Child Characteristics

The table below illustrates the direction and strength of associations between correct vaccination status and child characteristics.

##### B.2.3. Vaccination Status and Mothers’ Knowledge of Vaccination Schedule and VPDs

In the above table, children whose mothers have a fairly good level of knowledge about vaccine-preventable diseases were twice as likely to have a correct vaccination status compared to those born to mothers with poor knowledge levels.

### A. Multivariate Analysis (Final Logistic Model)

In order to determine the variables associated with correct vaccination status, variables with a p-value less than or equal to 20% in univariate analysis were introduced into the multivariate regression model. The table below provides the factors that remained significantly associated with correct vaccination status after modeling.

#### 1. Factors Associated with Correct Vaccination

The variables that remained statistically significant in the final model were: husband’s education level, household standard of living, distance from residence to vaccination facility, and mothers’ knowledge level of vaccine-preventable diseases. Adjusted for other variables:

- Children born to fathers with primary education are 15 times more likely to be correctly vaccinated than children born to fathers with no education [OR = 15.41]; education helps improve an individual’s healthcare-seeking behavior.
- Children from households with high standard of living are 10 times more likely to have complete vaccination [OR = 10.23] compared to those from households with low standard of living.
- Children born to mothers who perceive distance to the healthcare facility as a problem are 0.12 times likely to receive correct vaccination than children of mothers who don’t perceive distance as a problem [OR = 0.12].
- Based on the level of knowledge about vaccine-preventable diseases, children of mothers with fairly good knowledge are 2 times more likely to have correct immunization [OR = 1.73] compared to children of mothers with poor knowledge level.

#### 2. Model Validity

The model’s ability to correctly classify observations was evaluated using the ROC curve.

**Figure 1:**
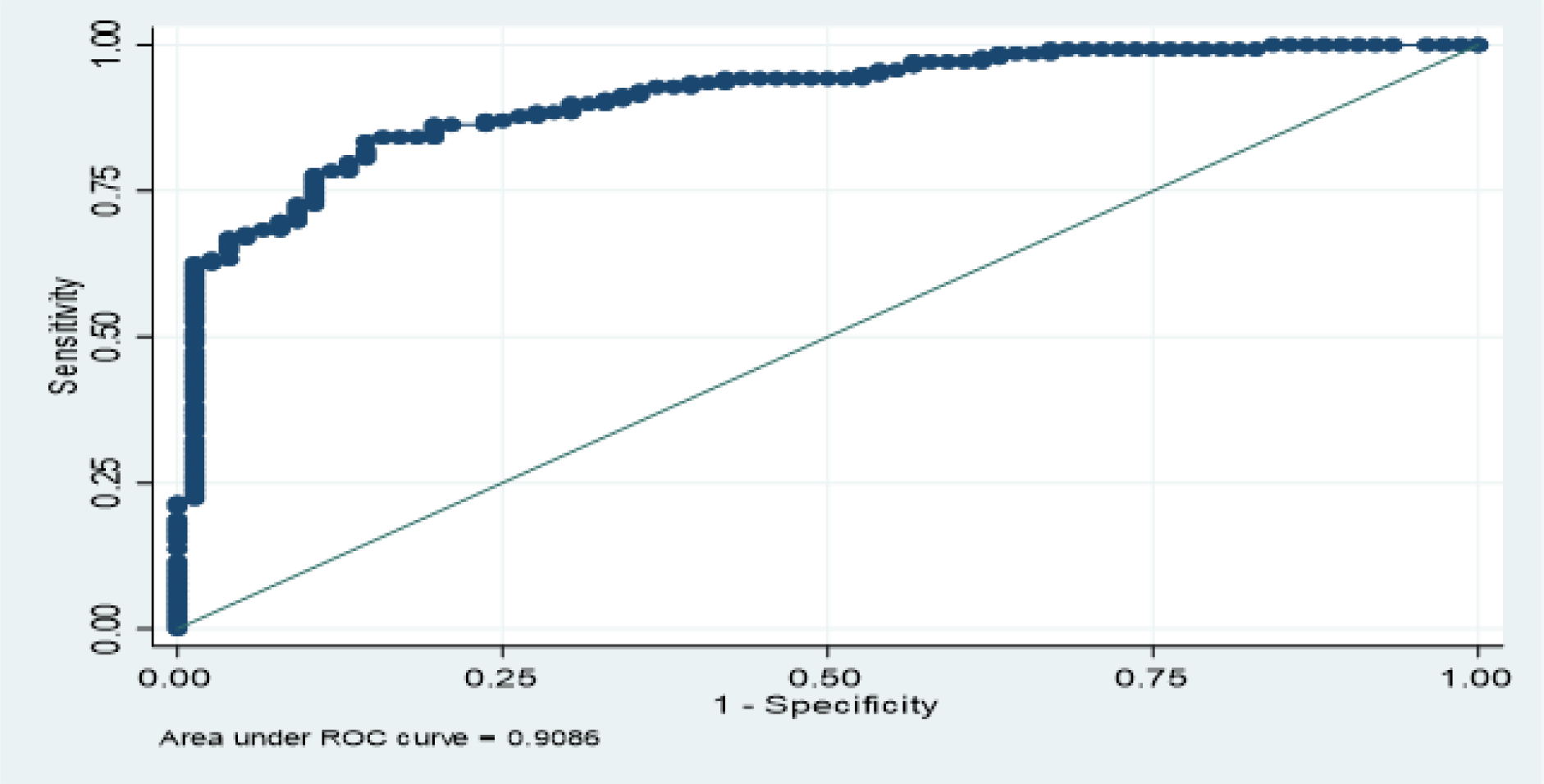
Model Validity.

The area under the curve is 0.9086. The model’s ability to correctly classify observations is 90%.

#### 3. Model Calibration

The Hosmer-Lemeshow test was applied to test the following hypotheses:

- H0: Theoretical probabilities are close to observed ones (calibrated model)
- H1: Theoretical probabilities are different from observed ones (non-calibrated model) The p-value of the Lemeshow test is 0.6697 (value > 0.05), indicating that the model is calibrated

## IV. Discussion

This study was conducted to identify predictors influencing correct vaccination of children aged 24 to 48 months hospitalized in 4 national hospitals in Burundi. The discussion of our work’s results focuses on three points:

- Sample characteristics.
- Comparison of our study results with those of other studies.
- Limitations of our study.

### IV.1. Sample Characteristics

This study aimed to identify factors influencing the vaccination status of children hospitalized in the four national hospitals in the city of Bujumbura. The percentage of children correctly immunized was 64.81%. Among the included children, we had 56.94% boys and 43.06% girls, resulting in a sex ratio of 1.32%. This result is slightly similar to those of Nigeria, which found 50.4% boys (**10**), and those of Zhang Xinyi et al. in 2018 in China and Cameroon (**11**) with 50% boys. These results do not reflect national statistical data, where there are generally more girls than boys according to the EDS 2016-2017. The majority of children (57.41%) were the first or second child of their mother. First-born children represented 31.48%. These results are similar to those of the series by Adedire et al. in Nigeria, where first-borns represented 31.3%, and about 60% of children occupied at least the 2nd rank in the birth order (**10**). Our results could testify to a good birth control policy through good awareness of Family Planning in Burundi.

### IV.2. Factors Associated with Correct Vaccination Status of Children

Logistic regression was used to identify predictors of complete vaccination among children aged 24 to 48 months. The results show that the husband’s level of education, household wealth, distance from home to vaccination center, and mothers’ knowledge of vaccine-preventable diseases were significantly associated with correct child vaccination. The father’s level of education had a significant positive influence on the likelihood of the child being correctly vaccinated. Children born to fathers with primary education are 15 times more likely to be correctly vaccinated [OR = 15.41] than children born to fathers with no education. Education helps improve an individual’s health service utilization behavior. This conclusion is consistent with other literature (**12–14**), which found education to be a significant predictor of vaccination completeness. In this study, it was also found that household wealth was positively associated with correct child vaccination. This means that the higher the household wealth, the more likely the child is to be correctly immunized. Children from wealthy households are 10 times more likely to have correct vaccination [OR = 10.23] compared to children from low-income households. Several studies have found a real relationship between wealth and vaccination status (Devasenapathy N and Ghosh Jerath S in 2016) (**15**). Children from richer households may be more likely to have correct vaccination status and receive missing vaccine doses when they attend a health facility than children from poor households. This may also be because children living in poor households have difficulty being reached by health workers, and parents may also struggle to access health facilities compared to children from wealthy households. However, in the study conducted by Rahman M, Obaida-Nasrin S in 2010 in Bangladesh (**12**), they found no association between wealth status and complete child vaccination.

Several studies have used education level and wealth as indicators of vaccination status. For example, a study conducted in the United States showed similar conclusions for these two factors, i.e., low education level and low socioeconomic status were both associated with incomplete or delayed vaccination (**16**). It corroborates a systematic review (studies conducted in low- and middle-income countries) that showed that low education level and low socioeconomic status were often strongly correlated and associated with incorrect vaccination (**17**). A significant association was found between distance to health facility and correct child vaccination in this study. Distance from the health facility strongly influenced a child’s vaccination status. Children born to mothers who reported that distance to the health facility was a problem were 0.12 times more likely to receive correct vaccination than children whose mothers reported that distance was not a problem [OR = 0.12].

In previous studies conducted in other developing countries, distance to primary health care facilities significantly predicted vaccination status (**12, 13**). The same results were observed in the study conducted in Kenya by Mutua et al. in 2011 (**14**). This may be because parents may not want to travel long distances due to the regular absence of health workers or unavailability of vaccines at the health facility. Another explanation could be that the visibility of the health facility could serve as a reminder to the parent. Similar results were also found in the study conducted in Tanzania (**18**). Although most medicines are provided free of charge, indirect costs related to transportation may explain the difference in vaccination status between those living closer and farther from the vaccination facility.

The level of knowledge of vaccine-preventable diseases in our study series was significantly associated with vaccination status; in our study, children of mothers with fairly good knowledge were twice as likely to have correct immunization [OR = 1.73] compared to children of mothers with poor knowledge of vaccine-preventable diseases. Our results are consistent with those found in Nigeria, which show that the impact of mothers’ knowledge level of vaccine-preventable diseases was positive on the level achieved by child vaccination coverage (**19**).

### IV.3. Study Limitations

Our study was conducted in four national hospitals in the city of Bujumbura. Although these hospitals are national referral structures, the non-inclusion of non-sick children; children from other facilities limits the possibility of generalizing the results obtained in this study. The systematic exclusion of certain children may be up-to-date but not having vaccination cards constituted a significant limitation.

## V. Conclusions

For our study, only 64.81% of children are correctly vaccinated according to the PEV vaccination schedule. The prevalence of correct vaccination coverage among children remains low in the study sample. Our study identified various factors influencing this prevalence, including the household head’s education level, household wealth, distance between residence and vaccination facility, and mothers’ knowledge of vaccine-preventable diseases. It is therefore important to strengthen health education within the population and improve PEV activity offerings in peripheral health facilities, especially when the country is willing to introduce new vaccines into the PEV to reduce morbidity and mortality from vaccine-preventable diseases.

## Data Availability

The data used in this study is available upon request from the author “Felix Niyongabo”. Interested parties can contact the author at.

## REFERENCES

1. Immunization coverage [Internet]. [cited 2020 May 21]. Available from: https://www.who.int/news-room/fact-sheets/detail/immunization-coverage

2. Rapport d’évaluation du plan d’action mondial pour les vaccins [Internet]. 2014. 1-36 p. Available from : www.who.int/vaccine/SAGE_DoV_GVAP_Assessment_report_2014_French.pdf

3. Omer SB, Orenstein WA. Vaccine Refusal, Mandatory Immunization, and the Risks of Vaccine-Preventable Diseases. The New England Journal of Medicine. 2009;8.

4. Immunization_Summary_2013.pdf [Internet]. [cited 2020 May 25]. Available from: https://www.who.int/immunization/monitoring_surveillance/Immunization_Summary_2013.pdf?ua=1

5. Jm G, Sr N, Kj N, Sj H, Mf D, Nm W, et al. A population-based cohort study of undervaccination in 8 managed care organizations across the United States. JAMA Pediatr. 2013 Mar 1;167(3):274–81.

6. Gowda C, Dempsey AF. The rise (and fall?) of parental vaccine hesitancy. Hum Vaccin Immunother. 2013 Aug;9(8):1755–62.

7. MinisterodellaSalute. Calendario Vaccinale. Available from: http://www.salute.gov.it/portale/vaccinazioni/dettaglioContenutiVaccinazioni.jsp?lingua=italiano&id=4829&area=vaccinazioni&menu=vuoto. - Search Results [Internet]. PubMed.

8. Bobossi-Serengbé G, Fioboy R, Ndoyo J, Nakouné E. Les occasions manquées de vaccination chez les enfants de 0 à 11 mois à Bangui. J PédiatriePuériculture. déc 2014;27(6):289 93 - Search Results [Internet]. PubMed. [cited 2020 May 26].

9. Ministère à la Présidence chargé de la Bonne Gouvernance et du Plan [Burundi] (MPBGP), Ministère de la Santé Publique et de la Lutte contre le Sida [Burundi] (MSPLS), Institut de Statistiques et d’Études Économiques du Burundi (ISTEEBU), et ICF. 2017. Troisième Enquête Démographique et de Santé. Bujumbura, Burundi : ISTEEBU, MSPLS, et ICF.

10. Immunisation Coverage and Its Determinants Among Children Aged 12-23 Months in Atakumosa-west District, Osun State Nigeria: A Cross-Sectional Study - PubMed [Internet].

11. Zhang X, Syeda ZI, Jing Z, Xu Q, Sun L, Xu L, et al. Rural-urban disparity in category II vaccination among children under five years of age: evidence from a survey in Shandong, China. Int J Equity Health. 2018 Jun 22;17(1):87.

12. Rahman M, Obaida-Nasrin S et al. Factors affecting acceptance of complete immunisation coverage of children under five years in rural Bangladesh. Salud Publica Mex. 2010; 52(2):134–40. -

13. Jani JV, De Schacht C, Jani IV, Bjune G.Risk, et al. Factors for incomplete vaccination and missed opportunity for immunisation in rural Mozambique. BMC Public Health. 2008;8:161

14. Mutua M, Kimani-Murage E, Ngomi N, Ravn H, Mwaniki P, and Echoka E. Fully immunized child: coverage, timing and sequencing of routine immunization in an urban poor settlement in Nairobi, Kenya. Tropical Medicine and Health. 2016; 44(1):1–2.

15. Devasenapathy N, Ghosh Jerath S, Sharma S, et al. Determinants of childhood immunisation coverage in urban poor settlements of Delhi, India : a cross-sectional study. BMJ Open 2016 Aug 26;6(8):e013015

16. Suarez L, Simpson DM, Smith DR. The impact of public assistance factors on the immunization levels of children younger than 2 years. Am J Public Health 2007; 87(5):845–8.

17. Rainey JJ, Watkins M, Ryman TK, Sandhu P, Bo A, Banerjee K. Reasons related to non-vaccination and under-vaccination of children in low and middle income countries: findings from a systematic review of the published literature, Vaccine 2011;29(46):8215–21.

18. Le Polain de Waroux O, Schellenberg JR, Manzi F, Mrisho M, Shirima K, Mshinda H, Alonso P, Tanner M, Schellenberg DM. Timeliness and com-pleteness of vaccination and risk factors for low and late vaccine uptake in young children living in rural southern Tanzania. Int Health 2013; 5:139–47; PMID:24030114; 10.1093/inthealth/iht006 .

19. Odusanya OO, Alufohai EF, Meurice FP, Ahonkhai VI. Determinants of vaccination coverage in rural Nigeria. BMC Public Health. 2008 Nov 5;8:381

